# Microsimulation of SARS-CoV-2 Transmission in Society

**DOI:** 10.1101/2021.05.16.21257298

**Authors:** Anthony Roland Green, Daniel Keep, Ian Piper

## Abstract

The outbreak of the pandemic disease, COVID-19, has shown that the approaches by different countries has resulted in a range of infection rates through their societies. This has arisen from the varying personal behaviour and tactical use of lockdown strategies within each country. We report the use of microsimulation of a simulated community in Australia, using a discrete infection model within a community of residences, places of work and recreation to demonstrate the applicability of this method to both the current pandemic and to infection more generally. Simulations without any societal intervention on infection spread provided base simulations that could be compared with social and societal controls in the future and which were compared with the initial doubling times of country outbreaks across the world. Different population sizes were represented in some simulations and in other simulations the effects of either social distancing or the use of facial masks as personal behaviours was investigated within the community. Good agreement is found between the initial doubling times for several countries and the simulations that suggests that modelling infection as a collection of individual infections provides an alternative to current epidemiological models. The variation of the basic reproductive number, R_0_, with time and population size, suggests that one of the fundamentals assumptions in SIR type models is wrong, but varies according to the properties of the population being modelled. Investigation of the infection spread shows that the number of super-spreaders varies with the size of the population and occurs through contacts in clubs, supermarkets, schools and theatres where the source of infection is an employee and where there are high numbers of contacts. The simulations of individual control show that the benefits of social distancing or wearing masks is only fully realised where there is considerable compliance within society to these measures.

## 1 Introduction

The coronavirus disease, COVID-19, is caused by the transmission of the RNA virus Severe Acute Respiratory Syndrome Coronavirus 2 (SARS-CoV-2) which was first notified to the WHO on 31 December 2019 by Chinese authorities. At this time, a pneumonia of unknown cause had been detected in Wuhan. A report in the South China Morning Post [1] suggested that a 55-year-old man could have contracted the disease on 17 November and that by the time the WHO was notified of the new disease there were some 266 confirmed cases. No matter what the exact origins were, it has since been transmitted to most countries around the world.

## 2 Infection Model

The infection model used in this study is a seven-state model for each individual. The individual times spent in each compartment were based on the WHO report [2] which detailed preliminary infection parameters and recovery information from China. The infection model is shown diagrammatically in Figure 1. In the simulation all individuals start susceptible to the disease, Covid-19. The simulation is started by one infected person who is injected into the community.

**Figure 1.**
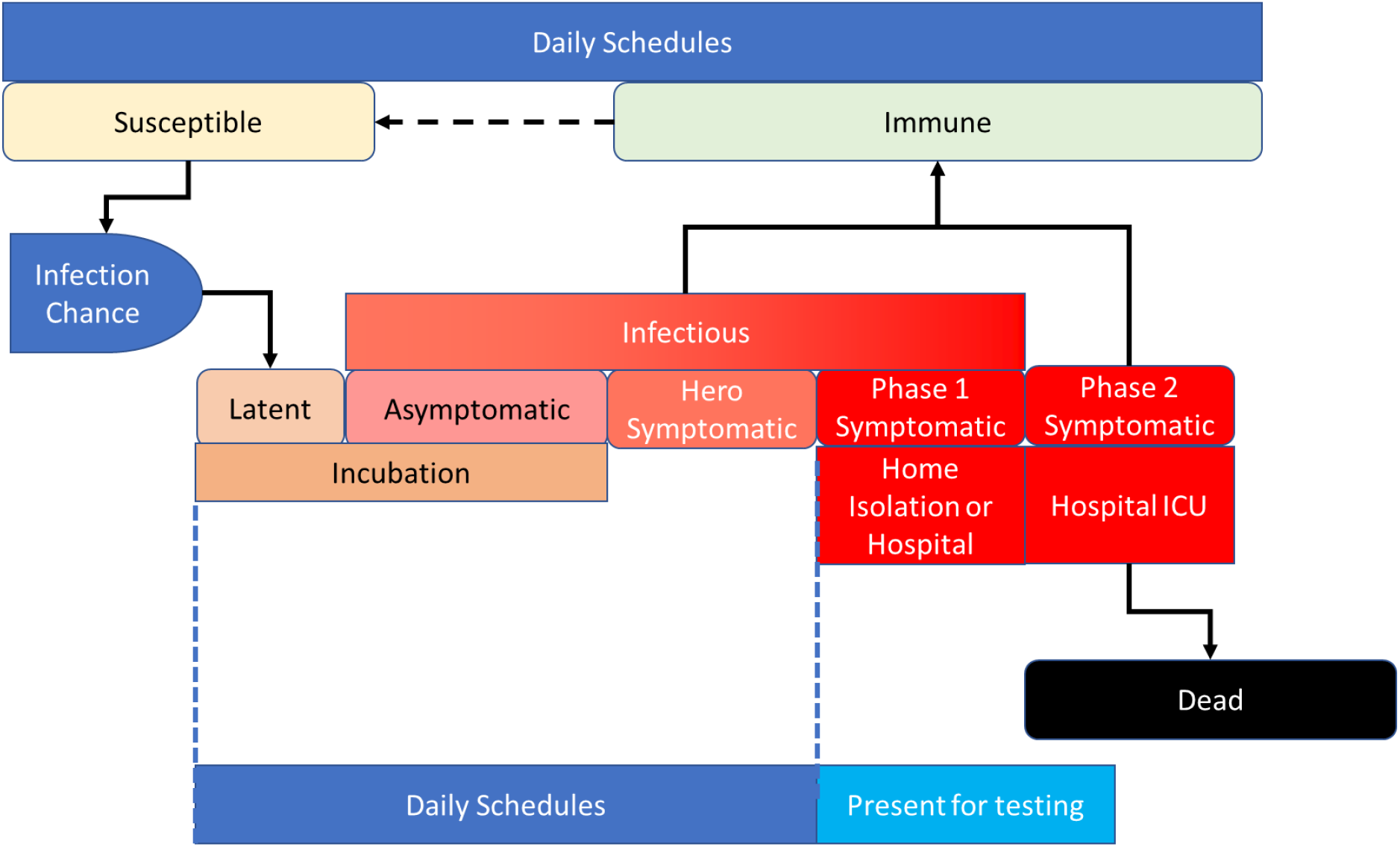
Schematic of the infection model used for both Covid-19 and an influenza like virus.

A second infection, to provide a mechanism for generating negative testing results, was not tested in the simulations presented in this paper. In NSW, the government asked that only those presenting with symptoms were to be tested. Only those that came back positive become confirmed cases. The simulation emulates this. All individuals present for testing on becoming symptomatic although this is delayed if the individual has a significant hero phase where the person knows they are symptomatic but continues their normal routine. Individuals who test negative for the SARS-CoV-2 virus continue their normal daily schedules. Individuals who test positive for covid-19 are home or hospital isolated until recovery and are counted as confirmed cases.

The susceptible population are delayed from becoming latent to either disease by an infection chance in the presence of an infectious individual in a location (cell). The equation for infection in a cell for each timestep is given by:

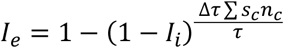

Where *I*_*e*_ is the chance of infection of the individual within a cell, *I*_*i*_ is the individual’s infection chance, n_c_, is the number of infectids in the cell, Δτ is the simulation timestep and τ is the average infectious time. *S*_*c*_ is the infection strength associated with infected *n*_*c*_. This parameter takes account of the variation in viral load coming from each infected individual but was set to 1 in all simulations presented here.

The latent and asymptomatic phases define the incubation period for the individual. In about 20% of cases the asymptomatic period is resolved directly to the immune state without going through a noticeable symptomatic phase and hence they do not get tested. The others progress to a hero symptomatic period which represents the onset of noticeable symptoms, but the individual ignores or conceals them and does not present for testing. At some point the individual enters the symptomatic phase 1 period where the individual presents for a covid-19 test. They are either sent home to isolate or are isolated in hospital if comorbid until their results come through. The delays in testing are built into the testing model. If Infected they continue home isolation or ward isolation. Those that go on to develop phase 2 symptoms are queued for a hospital ward, then ICU and then ventilator depending on their susceptibility to more severe outcomes. This part of the model has not been assessed in this paper. The individuals will then recover or die. Those that die are moved to a morgue to free up resources for other individuals. Those that recover are deemed to be immune and resume their daily schedules.

The infection parameters used in Table 1 have been derived from the WHO report [2].

**Table 1.** Global Infection parameters. Individual infection parameters are sampled from the distribution or via a random number comparing with a fraction between 0 and 1.

| Infection Parameter | Type | $\mu, \sigma$ or value | Comment |
| --- | --- | --- | --- |
| Incubation Interval | Lognormal distribution | 4.81, 0.042 |  |
| Symptomatic Interval | Normal distribution | 0.2, 0.01 | Individuals who become symptomatic. |
| Asymptomatic interval | Normal distribution | 5.47, 0.08 | Individuals who remain asymptomatic. |
| Asymptomatic cohort | Fraction | 0.2 | Population who remain asymptomatic. |
| Infection chance | Lognormal distribution | - | Varied in study. |
| Infection strengths: |  |  |  |
| Adult | Normal distribution | 0.9, 0.02 | Initial data suggested that children were not as prone to infection |
| Child | Normal distribution | 0.13, 0.1 |  |
| Health worker | Normal distribution | 0.25, 0.1 | Wearing effective PPE. |
| Hero Interval | Uniform distribution | 1 to 3 |  |
| Hero Cohort | Fraction | 0.11 |  |
| Symptomatic intervals: |  |  |  |
| Mild to recovery | Normal distribution | 5.47, 0.11 |  |
| Mild to severe | Normal distribution | 4.45, 0.11 |  |
| Severe to recovery | Normal distribution | 6.57, 0.21 |  |
| Severe to death | Normal distribution | 6.53, 0.24 |  |
| Symptomatic severity: |  |  |  |
| Mildly infected child | Fraction | 0.973 |  |
| Severely infected child | Fraction | 0.025 |  |
| Critical infected child | Fraction | 0.002 |  |
| Ventilated infected child | Fraction | 0.0015 |  |
| Mildly infected adult | Fraction | 0.8 |  |
| Severely infected adult | Fraction | 0.139 |  |
| Critical infected adult | Fraction | 0.061 |  |
| Ventilated infected adult | Fraction | 0.046 |  |
| Recovery Cohort | Fraction | 1 |  |

## 3 Community Structure

The community locations consist of two parts; residential locations based on the demographics from the 2016 census for the Nepean Health District, community locations for work, shopping, recreation and religion.[3] Table 2 contains the number of workplaces and different locations in the simulation [3].

**Table 2.** Locations used within the community. These have been assembled from references [3,4]

| Type | Location | Number | Comment |
| --- | --- | --- | --- |
| *Residential:* | Residences | 121465 | Does not include empty properties. |
| *Recreation:* | Park fraction | 0.000014 | Recreational locations were calculated from the fraction of the population in the simulation. |
|  | Stadia fraction | 0.000013 |  |
|  | Sports fraction | 0.00053 |  |
|  | Swimming fraction | 0.000065 |  |
|  | Religion fraction | 0.000506 |  |
| *Workplaces:* |  |  | Other places are defined from the number in the district population |
| Education | Tertiary | 9 |  |
|  | High schools | 51 |  |
|  | Primary schools | 116 |  |
|  | Kindergartens | 163 |  |
| Health | COVID-19 hospitals | 1 |  |
|  | Hospitals | 9 |  |
|  | F-practices | 102 |  |
|  | G-Practices | 193 |  |
|  | Diagnostic collection | 193 |  |
|  | Labs | 5 |  |
| Entertainment | Theatres | 9 |  |
|  | Cinemas | 14 |  |
|  | Concert halls | 23 |  |
|  | Clubs | 130 |  |
| Eateries | Restaurants | 391 |  |
|  | Hotels | 65 |  |
|  | Cafes | 391 |  |
|  | Takeaways | 391 |  |
| Food and Grocery | Superstores | 12 |  |
|  | Supermarkets | 30 |  |
|  | Groceries | 135 |  |
|  | General stores | 666 |  |
| Retail | Megastores | 12 |  |
|  | Large shops | 30 |  |
|  | Medium shops | 135 |  |
|  | Small shops | 666 |  |
| General | Work-1 | 121 |  |
|  | Work-2 | 302 |  |
|  | Work-3 | 1358 |  |
|  | Work-4 | 6670 |  |
|  | Work-5 | 13651 |  |

The community consists of six cohorts of people: infants (0-4-year-olds), primary (5-14-year-olds), secondary (15-24-year-olds), full-time employed, part-time employed, and unemployed [4]. The fraction of infants that go to preschool or equivalent outside the parent’s home are given schedules for the days on which they attend. The remainder are assumed to remain at home irrespective of where the parents are located. The primary cohort either go to primary school or secondary school. It is assumed that this cohort does not work. The secondary cohort go to secondary school, TAFE college or university. A sizable fraction is no longer in full time education and are added to full-time or part-time work of the adult population over the age of 24. A fraction of those who are still in full- or part-time education have part-time jobs. These attract specific schedules rather than those in the adult population.

The remaining three cohorts work or are unemployed [4]. Persons who have retired are added to the unemployed group. Table 3 is the breakdown of the different population cohorts used in the simulations.

**Table 3.** Population for the Nepean Health District based on the 2016 census data. [5,6]

| Cohort | Number | Comment |
| --- | --- | --- |
| District Population | 358637 | Total Population (Nepean Health District). |
| Infants | 28914 | Children 0-4 years of age. |
| Primary | 28981 | Children 5-14 years of age. |
| Secondary | 32785 | Students who attend secondary education only. |
| Full-time workers | 126905 | Includes a cohort of 20-24 year-olds who work full-time as well as attending tertiary education. |
| Part-time workers | 55937 | Includes students 15-19 and 20-24 who work part-time as well as attending secondary or tertiary education. |
| Unemployed/retired | 85115 | Unemployed, retirees and those identifying as at home. |
| Health worker cohort | 0.125 | Fraction of population who work in health care. |

## 4 Daily Schedules

All individuals are assigned a schedule for each day, with different schedules being chosen from a weighted random list for the cohort to which they belong. The weighting represents the chance that a particular schedule is followed. For example, a person working full-time might have a five-day week with two rest days. Consequently, they have a two in seven chance of not working. The process is more complicated than this as a full-time employee might be on shift work with a rotating roster of shifts and different starting times. Therefore, the ratios are different from just a 9-5 worker and two days off. Table 4 shows the options for schedules.

**Table 4.**
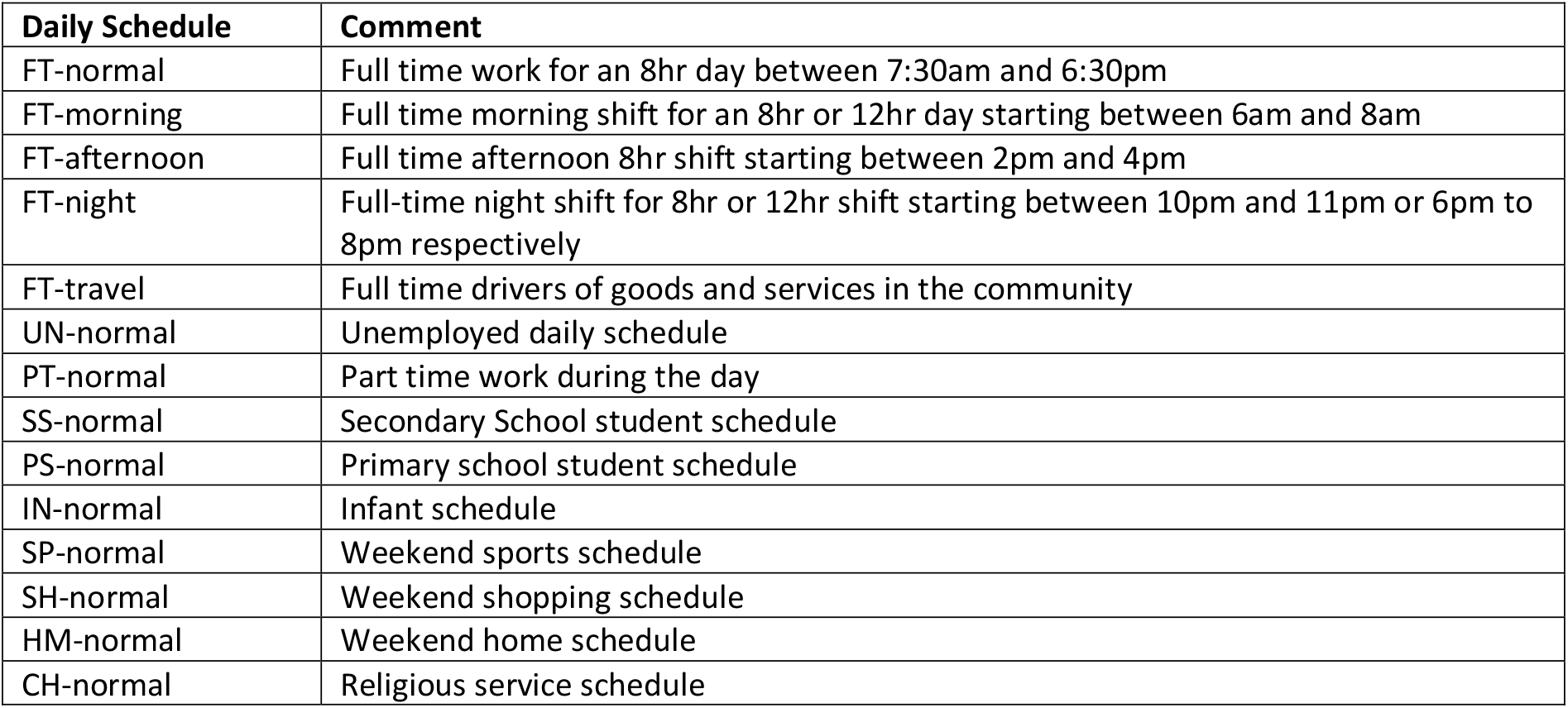
Daily Schedules for the population

| Daily Schedule | Comment |
| --- | --- |
| FT-normal | Full time work for an 8hr day between 7:30am and 6:30pm |
| FT-morning | Full time morning shift for an 8hr or 12hr day starting between 6am and 8am |
| FT-afternoon | Full time afternoon 8hr shift starting between 2pm and 4pm |
| FT-night | Full-time night shift for 8hr or 12hr shift starting between 10pm and 11pm or 6pm to 8pm respectively |
| FT-travel | Full time drivers of goods and services in the community |
| UN-normal | Unemployed daily schedule |
| PT-normal | Part time work during the day |
| SS-normal | Secondary School student schedule |
| PS-normal | Primary school student schedule |
| IN-normal | Infant schedule |
| SP-normal | Weekend sports schedule |
| SH-normal | Weekend shopping schedule |
| HM-normal | Weekend home schedule |
| CH-normal | Religious service schedule |

Each schedule assumes that a person moves to several different locations during the day. The individuals are ‘teleported’ between venues where the times at which consecutive movements occur being chosen at random within a given period. An example of these schedules is shown in Figure 2. A list of schedules is given in Table 4. The daily routine used for a given individual is randomly selected from weighted lists that represent the prevalence of different types of schedules in the community for the cohort type. For example, in the part-time cohort people may work but have schedules combining full-time, part-time and non-work (effectively unemployed) days. Consequently, the daily schedule is selected from a weighted list of PT-normal, FT-normal and UN-normal that represents proportions consistent with census data.

**Figure 2.**
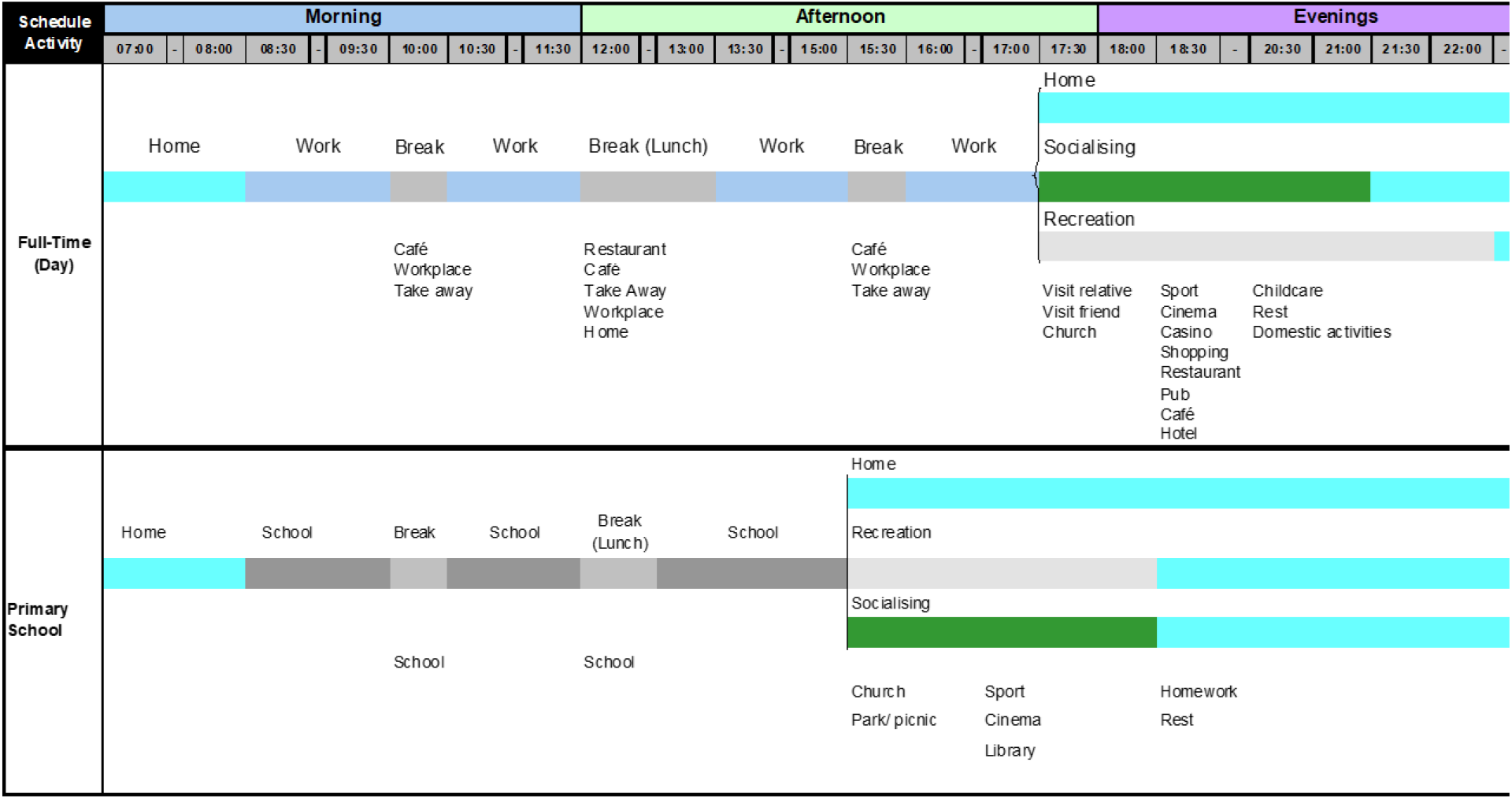
Two examples of scheduling; Top) A full-time day schedule for an individual employed full-time, Bottom) A schedule for a primary student during term.

## 5 Simulations

The Nepean Health District has a population of about 353000 persons. A fraction of this population was used in different simulations with the number of residences and community locations scaled accordingly. Simulation populations ranged from 700 to 212000 individuals to assess the effect of population size on how quickly infection will spread through a community; assuming there is no societal intervention. These can be compared with the spread in different countries, particularly where infection has spread through different size populations such as the initial outbreak in Northern Italy.

In all simulations, one index case is introduced into the community on 13 January, similar to the first case that occurred in NSW. The person has come from overseas and goes to stay with a relative in the community. A testing regime where people go to a doctor or hospital to be tested for Covid-19 is used to discriminate between the confirmed cases and those in the population who have covid-19 but have not been diagnosed giving estimates of the total infection spread in the community. Once tested, individuals go home or hospital where they wait for confirmation. Those that test positive isolate at home, move to a Covid-19 ward, ICU or ventilator depending on the severity of the case. Those that test negative return to a normal routine. In these simulations the resourcing for hospital facilities is not investigated and will be the subject of future investigations.

### 5.1 Comparison with other countries

Several simulation sets were undertaken in which the basic infection chance was varied. A single infected person was introduced into the community in each simulation.

18 simulations were undertaken for each infection chance. Those that showed propagation of the infection to others were averaged. Due to variation in time with the start of the outbreak, the growth was aligned from the day when five confirmed cases were observed in the individual runs. These averages are shown in Figure 3 for a population of 7200 and compares the variation with the change in the natural logarithm of the infection chance.

**Figure 3.**
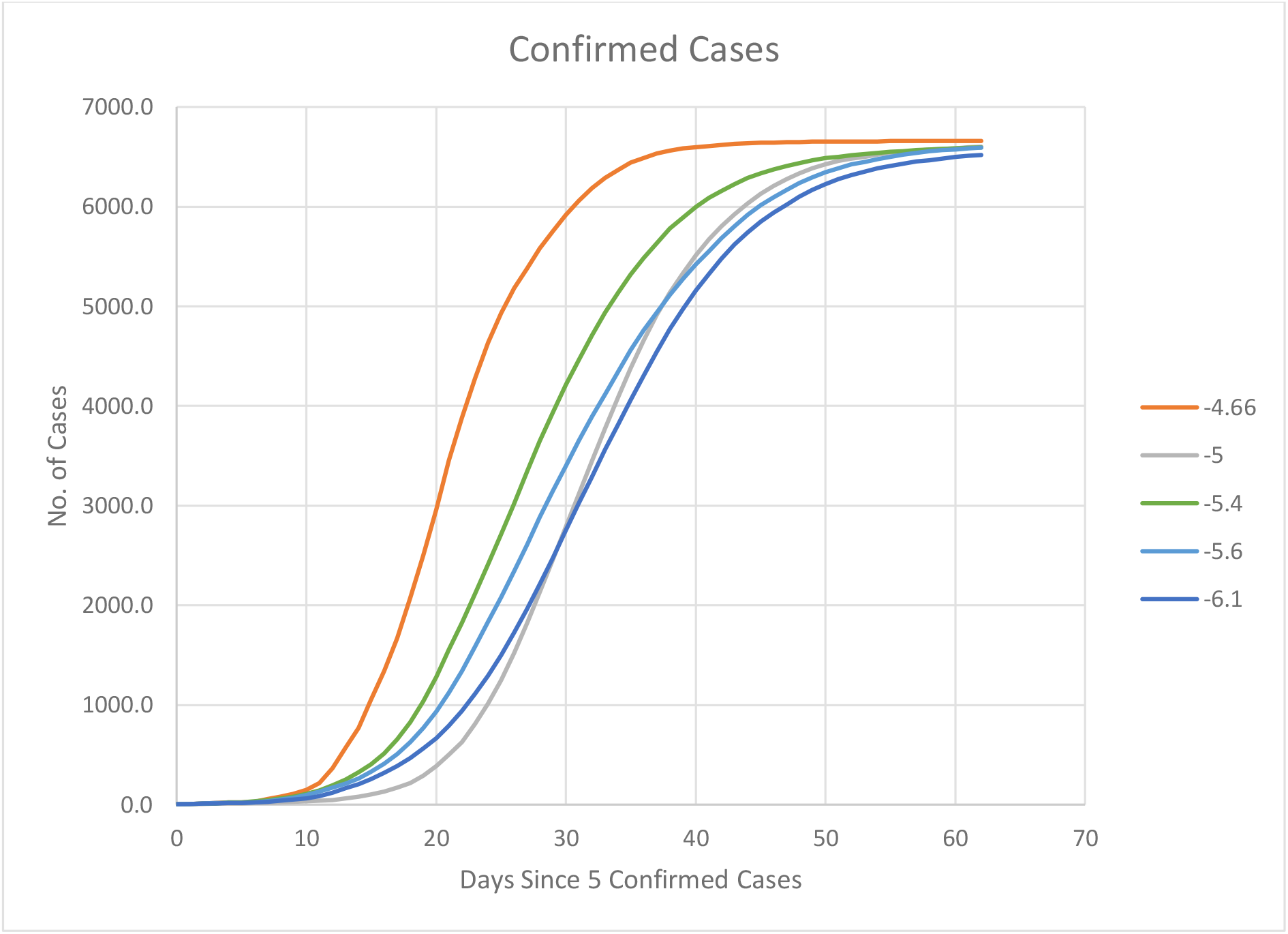
Variation with infection chance for a population of 7200. The legend represents Ln(infection-chance) used in the simulations.

The rates of growth from Figure 3, when expressed as the days to double the number of cases, is shown in Figure 4, together with the doubling times for several countries which did not act quickly on changing societal behaviour. Also shown is the doubling time for the NSW outbreak as a comparison. All doubling times were calculated from the rates of growth from 64 confirmed cases to 1024 confirmed cases.

**Figure 4.**
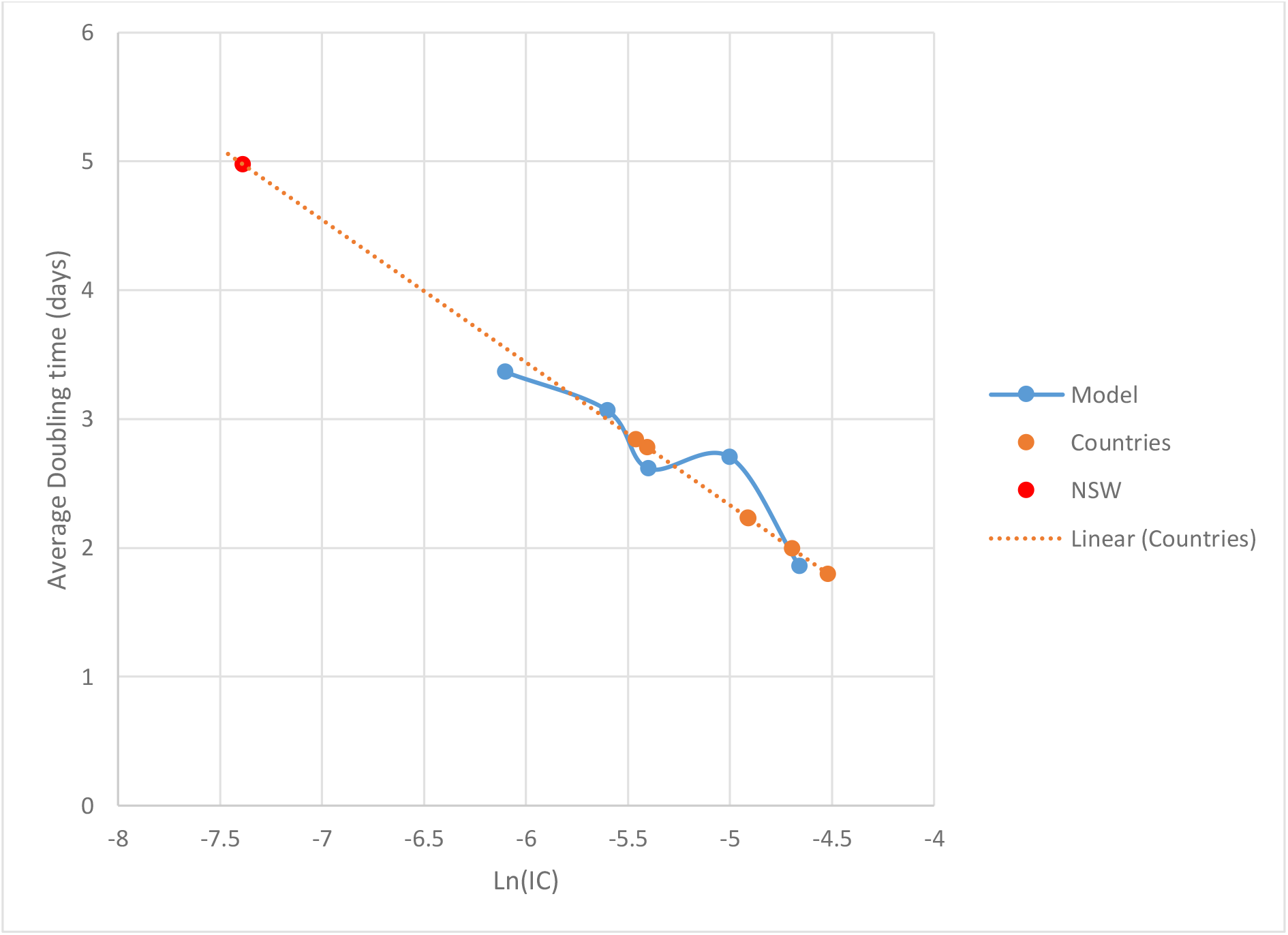
Comparison of Doubling Time with different country outbreaks. The doubling time was calculated from the growth rate between 64 and 1024 confirmed cases.

The growth rate was calculated from:

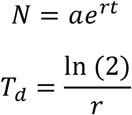

where *N* is the number of confirmed cases, *a* is a constant, *r* is the growth rate, *T*_*d*_ is the doubling time.

### Discussion

The country data was assembled from data available on the web [5] for the outbreaks in China, USA, Italy, France, UK, and Germany. The 64-1024 cases were used in the analysis because they would have occurred early in the pandemic, over a 7-to-15-day period, before most countries started to introduce government restrictions on movement.

All countries achieved 1024 confirmed cases within 55 days of their index case, and most occurred before day 42. The early data from Wuhan, China[2] suggested an R0 of 2.5 to 3 with an incubation period between 4.5 and 5.8 days with a mean of 5.1 days and the symptomatic phase for mild symptoms lasting 7 days on average. The delay between testing and results was at least 4 days at the start of the pandemic, no matter which country people were in, as the techniques had to be refined to bring the turn-around time down. As a result, confirmed cases would occur on average some 15 days after infection, assuming a delay of three days before being tested. 60 days therefore represents four generations of infection. Some governments may have started intervention earlier than this assumption but even if this was as early as 45 days then the infections statistics would not have altered significantly for the calculation of doubling time.

The data suggests that the infection chance for uncontrolled infection chance spread lies in the range 0.0022 to 0.009 per hour of contact with an infected person.

### 5.2 Variation with Population size

Several simulations investigated the effect of population size on the spread of infection, assuming no control over the movement of the population. The population size, along with the number of workplaces and residences, were calculated as a linear function of Nepean Health District population. A lower limit of 1 was set for hospitals, larger workplaces, clubs, cinemas and theatres, where the ratio was less than 1.

The average number of confirmed cases is shown in Figure 5. The results scale as expected with population. The graphs are self-similar with 85% of the population ending up being infected.

**Figure 5.**
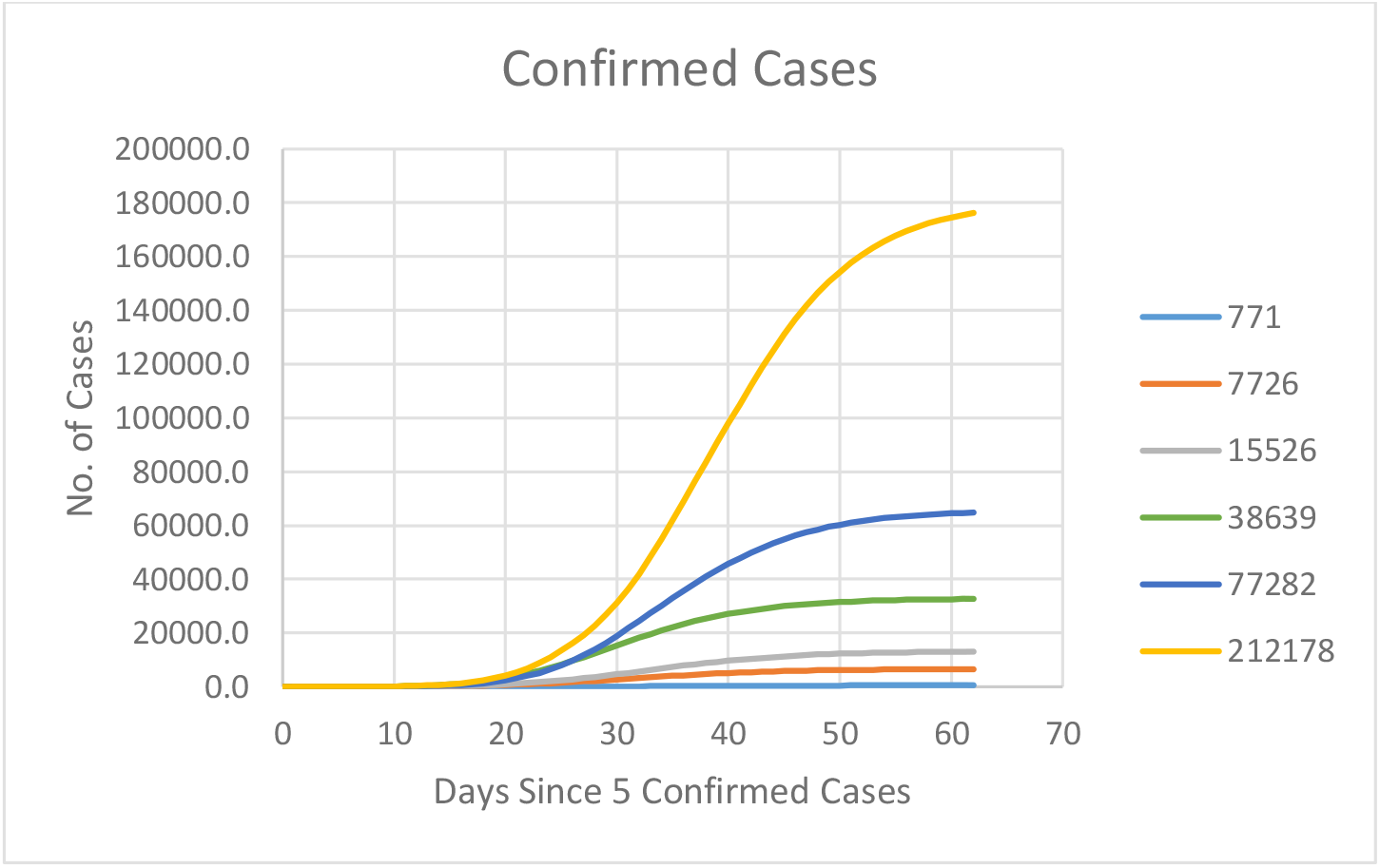
Variation in confirmed case numbers as a function of simulation population. The legend indicates the number of persons in the simulation.

Figure 6 shows a histogram of the fraction of the population infected from a person infecting more than one person. The majority of infected people cause no infection spread to other members of the population (60-66%). The histogram shows the presence of super-spreaders, that is where one individual infects many others. The inset shows the fraction of infections caused by super-spreaders in decade quanta for each population of total infections. The data suggests that, as the population increases, the fraction of super spreaders increases, although more simulations are required as the quantity will depend on the mixing model for the population and this has not been investigated in this work. The super-spreaders in the majority of cases are associated with workers in locations where people congregate such as clubs, theatres and schools or are schoolchildren.

**Figure 6.**
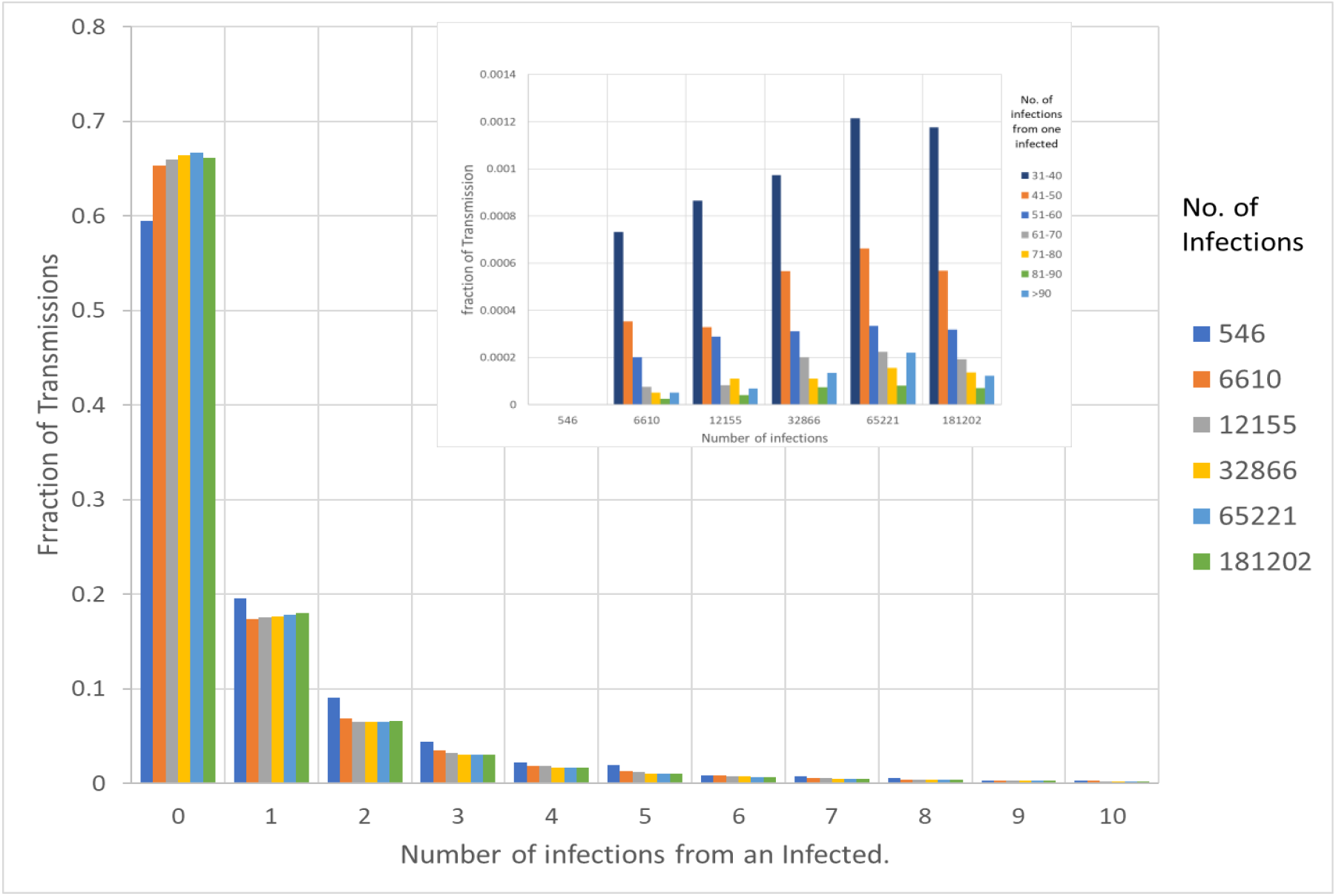
Histogram of transmission. The legend on the main diagram shows the total number of infections in each population simulation group. The legend in the inset indicates the numbers infected from super-spreaders in the community.

The Reproduction number, R_0_, is defined as the average number of infections caused by an infected person. Figure 7 shows the variation in R_0_ as a function of time and as a function of population. Higher maxima R_0_ values occur with higher populations.

**Figure 7.**
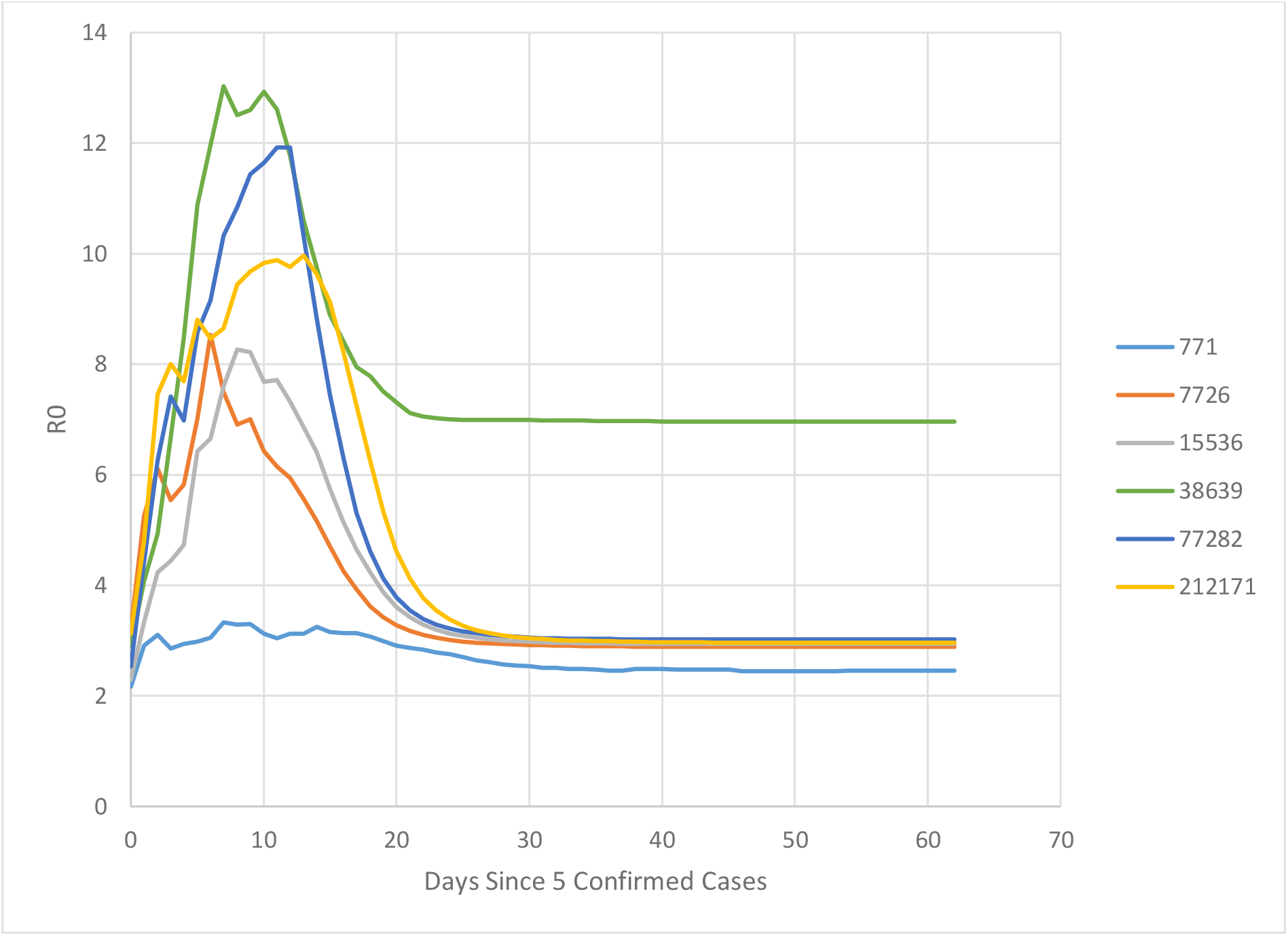
Reproductive number, R_0_, variance with time for different sizes of populations.

## Discussion

The results demonstrate that epidemiological models based on a constant reproductive number will overpredict the length of an infection wave as the reproductive number only tends towards a constant value later in the infection wave and well after the peak value occurs. The peak values seem to occur when 15% to 30% of the population have been infected.

The high reproductive numbers occur because of the presence of super-spreaders. It was notable that some of these were in primary and secondary schools being caused by children, despite their having a lower infection chance in the infection model than adults. The other people causing super-spreading work at clubs, theatres and supermarkets. As examples, there was only one super-spreader above 20 in a population of 771 people, where 26 people were infected at a club by one worker. In a population of 212171 people, there were examples of 1 schoolteacher infecting 130 high school students in one simulation, and people at home, in a stadium and at the theatre in another simulation.

### 5.3 Social distancing and mask wearing individual behaviours

Several test simulations were undertaken to gauge the effect of individual behaviour on infection rate. The social distancing and mask wearing behaviour are two inherent controls that individuals can undertake. Social distancing is reflexive (if I am distanced from you, then you are distanced from me) where mask wearing is not (my mask protects you to a different degree from the protection it affords me). The simple approach taken studies the effects of one or other of this type of control as the equation used is similar. This study does not examine both measures taken together, and this would require further investigation.

Two parameters were used to model behaviour; the fraction of the population who complied with social distancing or mask wearing, and the effectiveness of the individual control. Each individual either complied with the control being investigated or did not. The effectiveness of the control measure in the compliant population was modelled according to the following equation.

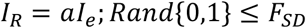

Where *I*_*R*_ is the reduced infection chance, *a* is the reduction in infectiousness of the control and *F*_*SD*_ is the fraction of the population who are compliant in the community.

Three series were used to investigate the variation of compliance in the population to either social distancing or mask wearing, with the efficacy of the action varied as a global parameter. Assuming that the population is 100% compliant in social distancing in venues or is mask wearing, Figure 8 indicates that as the efficiency increases to reduce the global infection chance, the rate of increase in infection spread decreases. All simulations were undertaken with populations of 771 people including the index case.

**Figure 8.**
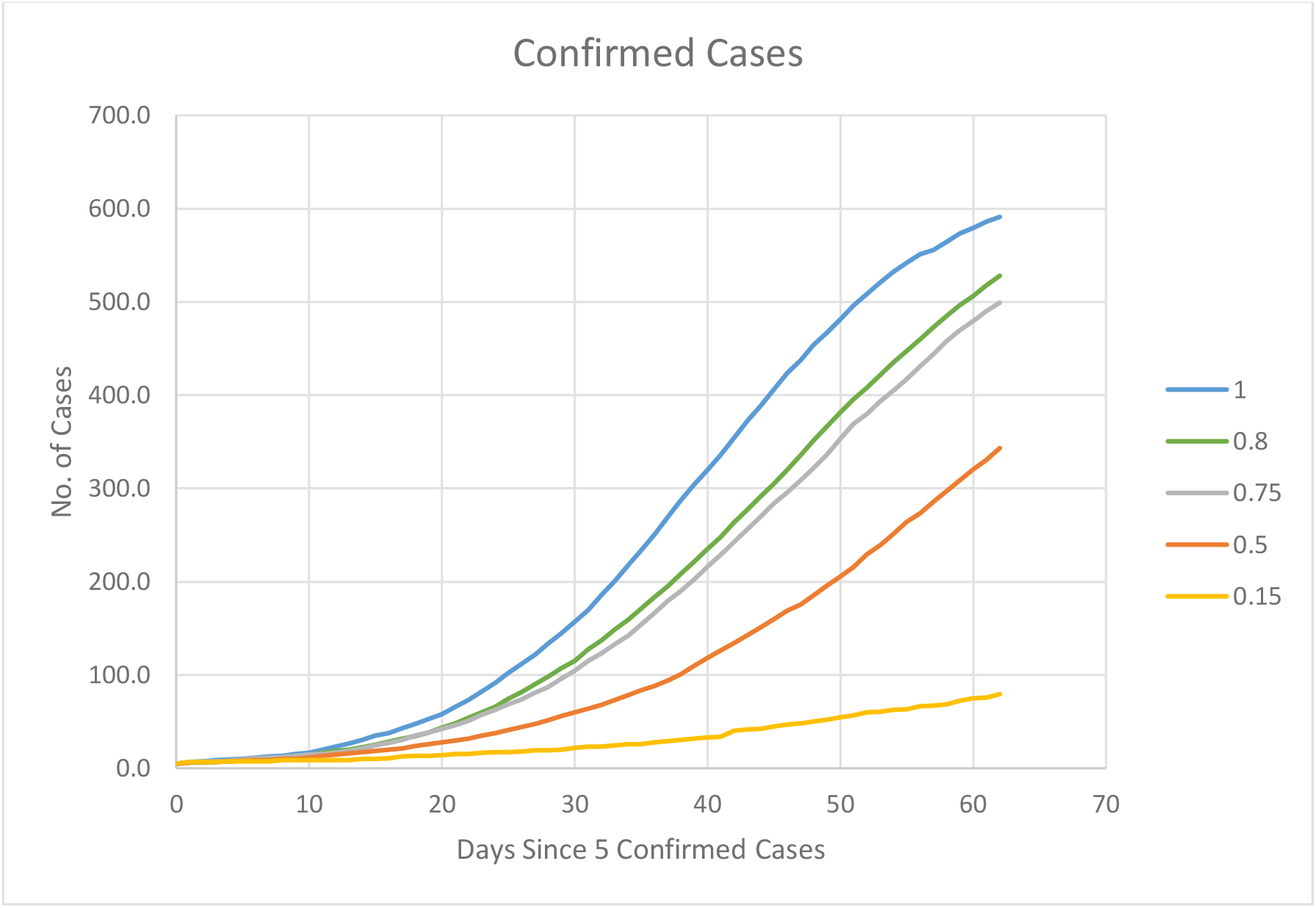
Effect of social distancing or mask wearing efficiency on a fully compliant population. The legend represents the inefficiency of the personal control. 1 therefore represents the zero efficiency as a control and 0 represents full efficiency as a control.

Figures 9 and 10 show the effect of relaxing the compliance to social distancing for efficiencies of social distancing of 85% and 30% respectively. It demonstrates that the as the efficiency in overall control decreases, compliance in the community is compressed into narrower bandwidths.

**Figure 9.**
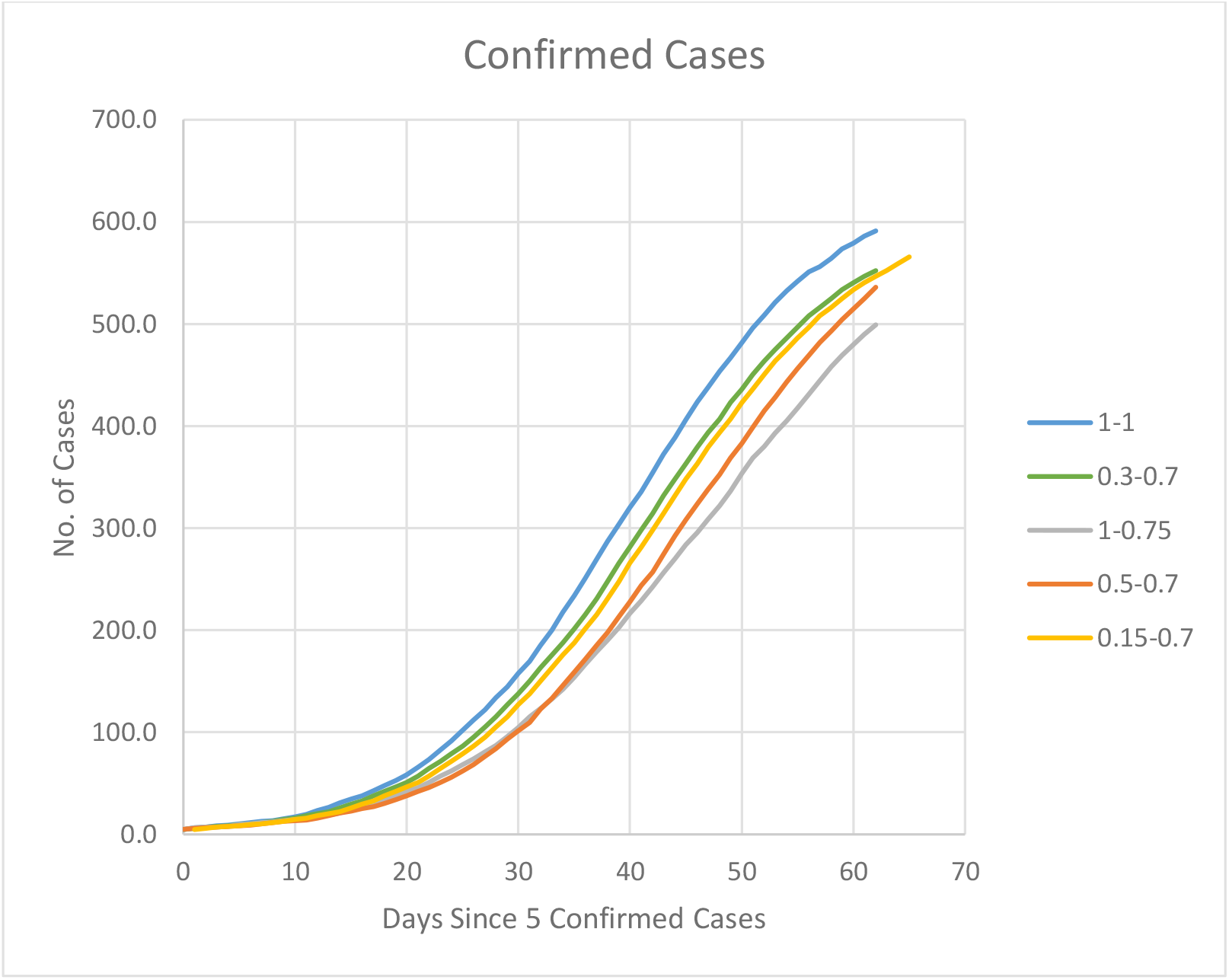
Effect of relaxing population compliance. The legend represents different degrees of compliance with an 85% efficiency of the control.

**Figure 10.**
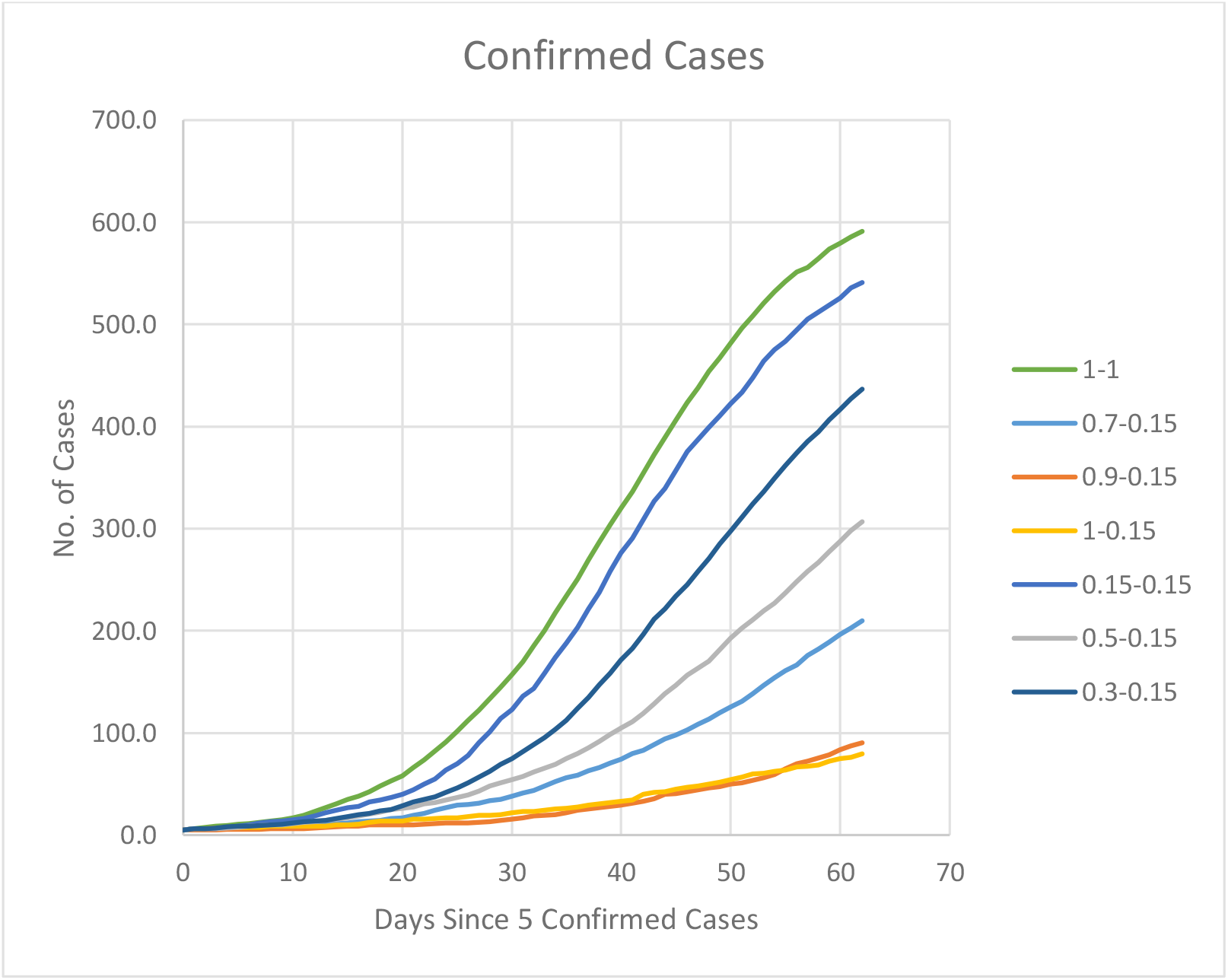
Effect of relaxing population compliance. The legend represents different degrees of compliance with an 30% efficiency of the control.

### Discussion

As individual behaviour becomes more compliant, either to social distancing or to mask wearing, the rate of disease spread decreases and the number of non-transmissions from the index case increases demonstrating ground truth in the techniques used.

## 6 Significance

Differential compartment models are commonly used in epidemiological studies [6-8] as an analytical tool for studying disease spread in society. They vary in complexity from the simple models to more complex models for the force of infection. Fundamental problems lie in the assumptions concerning the average number of contacts per person per time and the population that is considered as being susceptible to infection. In many cases this is assumed to be the total population and thus overpredicts the expected number of cases.

Because social mixing is so important to the prediction of infection in society, stochastic models of the population have been developed with smaller groups that better mimic the behaviours in society. The set of differential equations, however, are related back to SIR type relationships [9,10] and infection is still solved based on group dynamics rather than the individual.

In other cases, the compartments that are set up to mimic subgroup exposures assume different R0 values for each compartment in order to get agreement with historical cases. [11]

In this study a full societal model of mixing and infection is presented. The paper demonstrates that it is viable to observe the patterns of infection spread with a simple model based on individual behaviour. Its potential value lies in the ability to assess the effectiveness of behavioural controls in society, whether based on individual preference or government edicts. A further benefit may come from relating viral loads of individuals to the ability to spread infection spread in the community.

Further work is envisioned. While this model has not been fully calibrated, further work will be undertaken to calibrate this model against outbreaks in other countries and Australian Data. Further work will assess the impact of closing different types of business and social compliance, as well as the impact of resource constraints on the public health system and how the increase in death in society could be mitigated with alternative strategies.

## Data Availability

All data used in this article is available

## References

1 Josephine Ma, Coronavirus: China’s first confirmed Covid-19 case traced back to November 17, South China Morning Post, 13 March, 2020, updated 14 March 2020.

2 Report of the WHO-China Joint Mission on Coronavirus Disease 2019 (COVID-19), 16-20 February 2020, World Health Organisation, downloaded from https://www.who.int/docs/default-source/coronaviruse/who-china-joint-mission-on-covid-19-final-report.pdf on 17 May 2020.

3 Australian Bureau of Standards, Statistical Area 2 2016 Census data: Population Statistics, Residence statistics Employment Statistics, Education Statistics, Transport and Vehicle Statistics, and business participation downloaded from https://www.abs.gov.au/census between September 2017 and June 2018.

4 Australian Bureau of Standards, Statistical Area 2 population estimates, downloaded from https://www.abs.gov.au/ on 14 October 2019.

5 https://www.worldometers.info/coronavirus/

6 Kermack WO, McKendrick AG (1927) A contribution to the mathematical theory of epidemics. Proceedings of the Royal Society of London Series A 115: 700–721.

7 Halloran, E. M. (2001) Epidemiologic methods for the study of infectious diseases. Oxford University Press

8 Mathews JD, McCaw CT, McVernon J, McBryde ES, McCaw JM (2007) A Biological Model for Influenza Transmission: Pandemic Planning Implications of Asymptomatic Infection and Immunity. PLoS ONE 2(11): e1220. doi:10.1371/journal.pone.0001220

9 Ferguson, N. M., D. A. T. Cummings, S. Cauchemez, C. Frazer, S. Riley, A. Meeyai, S. Iamsirithaworn, and D. S. Burke (2005, September). Strategies for containing an emerging influenza pandemic in south east asia. Nature 437(8), 209–214.

10 Chao DL, Halloran ME, Obenchain VJ, Longini IM Jr (2010) FluTE, a Publicly Available Stochastic Influenza Epidemic Simulation Model. PLoS Comput Biol 6(1): e1000656. https://doi.org/10.1371/journal.pcbi.1000656

11 Oliver M. Cliff, Nathan Harding, Mahendra Piraveenan, E. Yagmur Erten, Manoj Gambhir, and Mikhail Prokopenko, Investigating Spatiotemporal Dynamics and Synchrony of Influenza Epidemics in Australia: An Agent-Based Modelling Approach, arXiv:1806.02578v1, 7 June 2018.

